# Evaluating malaria elimination strategies in military forces in Cambodia: a cluster-randomized clinical trial comparing monthly prophylaxis with focused screening and treatment

**DOI:** 10.1101/2024.11.26.24318027

**Authors:** Mariusz Wojnarski, Sidhartha Chaudhury, Threechada Boonchan, Rathvicheth Bun, Soklyda Chann, Panita Gosi, Kin Soveasna, Sokhun Song, Nillawan Buathong, Mali Ittiverakul, Sabaithip Sriwichai, Montri Arsanok, Worachet Kuntawunginn, Piyaporn Saingam, Chaiyaporn Chaisatit, Alongkot Ponlawat, Thanyalak Fansiri, Pattaraporn Vanachayangkul, Boonsong Jaichapor, Muth Sinoun, Char Meng Chuor, Thay Kheangheng, Mary So, Elizabeth Wanja, Silas Davidson, Michele Spring, Huy Rekol, Lek Dysoley, Kong Saly, Jeffrey R. Livezey, Jessica T. Lin, Philip L. Smith, Prom Satharath, Jessica E. Manning, Somethy Sok, David L. Saunders

## Abstract

**Background:** Identifying effective malaria elimination strategies for remote forested regions in Southeast Asia is challenging given limited resources. In this study, two malaria elimination strategies were evaluated in partnership with the Royal Cambodian Armed Forces - monthly malaria prophylaxis (MMP) and focused screening and treatment (FSAT).

**Methods:** Eight primarily military clusters (1,050 volunteers total) along the Cambodian-Thai border were randomized to 3 months of MMP or FSAT with monthly malaria testing by RDT, PCR, and microscopy for six months. Clusters were sub-randomized to permethrin treated (ITU) or sham water-treated clothing (sITU). Volunteers in MMP clusters were given three full monthly dihydroartemisinin-piperaquine (DP) treatment courses with 12 weekly 22.5mg primaquine. Volunteers in FSAT clusters were treated with appropriate first-line antimalarials if malaria-positive by microscopy or PCR.

**Results:** *Pf* positivity in MMP clusters was reduced by 90% (10% at enrollment to 1% at 6 months; absolute risk reduction (ARR) 9%) at 6 months. However, 32% of *Pf* cases treated with DP as MMP at baseline recrudesced, requiring rescue treatment at 1 month with artesunate-mefloquine. *Pf* positivity in FSAT clusters declined 66% over 6 months (7.6% to 2.7%; ARR 4.9%). MMP reduced *Pv* positivity from 9% to 0% at 3 months, but *Pv* rebounded to 6.7% at 6 months. FSAT failed to significantly reduce *Pv* positivity during the study. The 22.5mg weekly primaquine MMP regimen was safe, even for the 15% of volunteers with G6PD-deficiency. Those wearing ITU had additional *Pv* parasitemia reductions compared to sITU in the FSAT but not MMP groups. PCR was more sensitive than microscopy and RDT for detecting both species.

**Conclusions:** MMP was safe, and superior to FSAT to reduce *Pf* and *Pv*, suggesting greater utility to achieve malaria elimination in Cambodia. Low dose (22.5mg) weekly primaquine was a safe adjunct in this setting, even for those with G6PD-deficiency. Permethrin-treated clothing further reduced *Pv* parasitemia for FSAT but not MMP. MMP may be more easily scaled to eliminate malaria. The military may provide substantial support for regional elimination efforts.

## Introduction

Intensive containment and control efforts have successfully reduced malaria prevalence in the Greater Mekong Subregion (GMS). However, multi-drug resistant (MDR) *Plasmodium falciparum (Pf)* remains endemic in border areas of Cambodia and Thailand, with confirmed spread to neighboring countries [1]. In Southeast Asia, declining efficacy of artemisinin combination therapies (ACT) has occurred within a few years of their introduction as first-line therapies [2–5]. Concerns that ‘untreatable’ malaria may emerge, combined with relatively low burden favors an aggressive approach to elimination in the region. This has been codified in commitments by ASEAN leaders in 2018 and the World Health Organization in 2022 to a goal of regional elimination by 2025 for *Pf* and by 2030 for *Plasmodium vivax* (*Pv*) [6–8]. Unfortunately, the spread of MDR *Pf* has outpaced development of new antimalarials, limiting pharmacological approaches to elimination [9]. Malaria elimination is also particularly challenging among mobile populations with occupational exposures occurring in forested areas to include the military [10, 11]. Populations residing or working in forested border areas carry a significant MDR malaria burden in the GMS [12, 13]. There is currently no consensus on how elimination will be achieved for hard-to-reach mobile populations disproportionately affected by malaria [10, 11]. Multiple approaches have been suggested, most requiring further evaluation [14].

Two approaches are typically proposed for malaria elimination in this region. The first is ‘mass drug administration’ (MDA) and the second is ‘focused screening and treatment’ (FSAT) where entire communities are evaluated and positive members receive treatment. MDA involves mandatory treatment ranging from focal areas of outbreak to entire endemic communities. While there have been a large number of MDA studies conducted over the past 70 years, a recent systematic review identified numerous challenges in comparing studies of varying designs. MDA studies are often complicated by a lack of well-defined objectives [15]. MDA if often thought to be the more effective approach because of lower costs, simplified logistics and better outcomes despite known limitations that includes potential added risk to those without malaria. Unless sustained, MDA has had limited effectiveness in practice among diverse settings [16].

Prior studies identified factors associated with MDA success. These include achieving at least 80% population coverage, directly observed treatment (DOT), strong community engagement, and the use of 8-aminoquinolines, particularly in *Pv* transmission settings [17, 18]. However, increasing prevalence of MDR *Pf* may compromise MDA effectiveness, and repeated administrations may be necessary in high transmission areas [19]. Finally, the use of compulsory anti-malarial treatment courses with associated side-effects in healthy individuals raised ethical concerns for MDA in low-transmission settings. Despite documented logistical challenges to implementation [20], an FSAT approach was thought to be a more ethical though less effective alternative [21]. The FSAT and MDA approaches have not been evaluated head-to-head in Cambodia, and there have been few studies conducted in hard-to-reach mobile populations, particularly the military [22]. Despite widespread use of malaria chemoprophylaxis by Western militaries deploying to malaria endemic areas, this approach has not been systematically evaluated nor widely adopted by militaries in Southeast Asia.

Diagnostic challenges also influence potential approaches. Rapid diagnostic tests (RDTs) are a practical option to screen large populations but low sensitivity limits their ability to identify individuals with subclinical infections and low parasitemia in areas approaching malaria elimination [19, 23, 24]. Significant numbers of asymptomatic cases may be missed in the FSAT approach unless more sensitive methods such as PCR are employed [23]. PCR offers greater sensitivity and specificity but is expensive, difficult to implement in austere locations, and does not detect the latent stages of *P. vivax* [25]. Even without PCR, the amount of laboratory support and follow-up required to perform adequate systematic FSAT on even a monthly basis is significant [20]. The other key diagnostic challenge is detecting G6PD-deficiency for those being administered 8-aminoquinolines. While several new G6PD tests have been field tested in Southeast Asia, the gold standard quantitative testing remains expensive and difficult to implement in austere environments [26].

Operational costs and complexities for various elimination approaches are poorly quantified and may be underestimated. The optimal approach has yet to be defined but is likely to include vector control measures in addition to drug therapy. Vector control measures such as indoor residual insecticide spraying (IRS) and the use of insecticide treated nets (ITNs) have been widely employed in Cambodia. However, the efficacy of vector interventions suitable for militaries, including use of insecticide-impregnated uniforms and various forms of personal and spatial repellants, remain untested in field settings, particularly against disease endpoints in clinical trials. The contribution of insecticide resistance to effectiveness also remains unclear [27]. The potential roles of militaries, thought to be a potent force for elimination, have yet to be evaluated [28].

The present study aimed to answer some of the key outstanding questions outlined above to define optimal approaches to malaria elimination. The US Army Medical Component of the Armed Forces Research Institute of Medical Sciences (AFRIMS) conducted a cluster-randomized interventional malaria elimination trial in partnership with the Royal Cambodian Armed Forces (RCAF) and National Center for Parasitology Entomology and Malaria Control (CNM) in Cambodia. The study aimed to quantify the relative safety and effectiveness of two major interventional approaches to malaria elimination in this region: (1) monthly malaria prophylaxis (MMP) using 3 monthly 3-day treatment courses of dihydroartemisinin-piperaquine (DP) and a weekly low dose of 22.5mg of primaquine, and (2) focused screening and treatment (FSAT) of only those found to be either microscopy or PCR malaria positive using the current first-line antimalarials recommended for *Pf* and *Pv* respectively. The additive effects of insecticide-treated uniforms (ITU) on malaria prevalence were also assessed by comparing to sham-treated uniforms or clothing (sITU) to prevent infectious mosquito bites.

## Materials and Methods

Details of the study protocol and methods have been previously published [29]. The overarching goal of this pilot elimination study was to increase Cambodian malaria control program capabilities to diagnose, prevent, and treat malaria with a focus on at-risk military personnel. To do so, a scalable military malaria elimination ‘unit of action’ was established at the provincial level. The Unit primarily relied on medical personnel stationed with volunteers to monitor for disease and provide DOT. The military elimination unit was staffed by 187 RCAF personnel trained and supplied by the AFRIMS-CNM research team. This team included 170 medics assigned to individual units responsible for day-to-day treatment and follow-up during the study. Medics were divided roughly evenly between clusters with centrally located administrative, laboratory and IT support staff.

The primary objective of the study was to compare the effectiveness of two elimination approaches. A cluster-randomized, open label interventional study was conducted in eight military residential clusters on the Cambodia-Thai border. Each cluster included approximately 120 individuals. The study aimed to determine feasibility to achieve significant reductions in malaria prevalence using a combination of available interventions. There were four clusters where reactive screening and treatment (FSAT) was employed. In FSAT clusters, malaria was confirmed by real-time PCR, with those found positive by PCR treated with currently recommended medications according to national treatment guidelines. The other four clusters received monthly malaria prophylaxis (MMP) with dihydroartemisinin-piperaquine (DP) and 12 weekly doses of low-dose primaquine (PQ) (see Study Interventions). Clusters were sub-stratified *a priori* based on low or high malaria transmission assessed through routine public health surveillance prior to the study (see Table 1). Clusters were further sub-randomized to wear of permethrin (ITU) or sham-treated (sITU) uniforms or clothing while in forested areas.

**Table 1.** Malaria elimination cluster characteristics. Composition of 8 geographically isolated clusters randomized in the study including volunteer demographics, interventional assignments, baseline malaria prevalence, G6PD-deficiency prevalence and primaquine treatments administered. Values are expressed as # (%) of those enrolled in each cluster in rows 6 through 15. The number screened represented the population of each cluster, with the number enrolled corresponding to intervention coverage (94% overall).

| Cluster | A | B | C | D | E | F | G | H | Total |
| --- | --- | --- | --- | --- | --- | --- | --- | --- | --- |
| **Treatment Group** | FSAT | MMP | MMP | FSAT | FSAT | FSAT | MMP | MMP |  |
| **Uniform Assignment** | sITU | sITU | ITU | ITU | ITU | sITU | sITU | ITU |  |
| **Baseline Endemicity** | High | High | High | High | Low | Low | Low | Low |  |
| **Number Screened** | 151 | 150 | 134 | 129 | 121 | 139 | 140 | 150 | 1,114 |
| **Number Enrolled** | 143 (95) | 142 (95) | 130 (97) | 125 (97) | 115 (95) | 133 (96) | 128 (91) | 134 (89) | 1,050 (94) |
| **Non-military** | 68 (48) | 61 (43) | 48 (37) | 43 (34) | 33 (29) | 0 (0) | 1 (1) | 0 (0) | 254 (24) |
| Children | 32 (22) | 32 (23) | 33 (25) | 13 (10) | 10 (9) | 0 (0) | 0 (0) | 0 (0) | 120 (11) |
| Civilian adults | 36 (25) | 29 (20) | 15 (12) | 30 (24) | 23 (20) | 0 (0) | 1 (1) | 0 (0) | 134 (13) |
| **Positive for Malaria on Screening by PCR** | 29 (20) | 29 (20) | 28 (21) | 17 (14) | 4 (3) | 3 (2) | 22 (17) | 13 (10) | 145 (14) |
| *Pf* infection | 23 (16) | 17 (13) | 12 (9) | 12 (10) | 2 (2) | 2 (1) | 11 (9) | 5 (4) | 84 (8) |
| *Pv* infection | 5 (3) | 11 (8) | 11 (8) | 5 (4) | 2 (2) | 1 (1) | 9 (7) | 8 (6) | 53 (5) |
| Mixed *Pf/Pv* infection | 1 (1) | 1 (1) | 5 (4) | 0 (0) | 0 (0) | 0 (0) | 2 (2) | 0 (0) | 9 (1) |
| **G6PD Deficiency WHO Class I-III** | 15 (10) | 17 (12) | 25 (19) | 15 (12) | 13 (11) | 25 (19) | 23 (18) | 27 (20) | 160 (15) |
| G6PD Deficiency Primaquine-treated | 5 (3) | 16 (11) | 25 (19) | 2 (2) | 2 (2) | 1 (1) | 22 (17) | 24 (18) | 97 (9.2%) |
| >10% Hemoglobin Drop at Day 3 | 0 (0) | 3 (2) | 6 (5) | 0 (0) | 1 (1) | 0 (0) | 3 (2) | 1 (1) | 14 (1.3%) |

### Study population

Inclusion criteria included: 1) Volunteers 18-65 years of age and their dependents > 2 years of age, to include military and eligible Cambodian civilians eligible for care at an RCAF facility; 2) Able to give informed consent/assent; 3) Residing in the selected study areas, 4) Available for monthly follow-up for the 6-month study duration; 5) Agreed not to seek initial outside medical care for febrile illness unless referred by the study team; and 6) Authorized by the local Commander to participate in the study if on active military duty.

Exclusion criteria included: 1) Known allergic reaction or contraindication to interventions to be used to include dihydroartemisinin-piperaquine, artesunate-mefloquine, or primaquine; 2) Pregnant or lactating females, and females of childbearing age unwilling to use acceptable forms of contraception during the study. Study assessments took place in RCAF facilities near the Cambodia-Thai border.

### Cluster randomization, allocation and blinding

The eight community clusters were randomized to either MMP or FSAT groups, using time and region-blocked randomization. This ensured that randomized clusters were geographically distinct, and that treatment (MMP vs. FSAT) and uniform conditions (ITU vs. sITU) were evenly distributed among high and low transmission areas. There were four clusters in high and four in low endemicity areas. Volunteers meeting enrollment criteria received either permethrin-treated uniforms (ITUs) or sham (water-treated) uniforms or civilian clothing as appropriate (sITUs). Clothing was treated according to cluster assignment in single-blind fashion with the volunteer, but not the investigators, blinded to assignment. Diagnostic microscopists were blinded to each other’s readings and to study arm assignments. The study design in shown in Figure 1.

**Figure 1.**
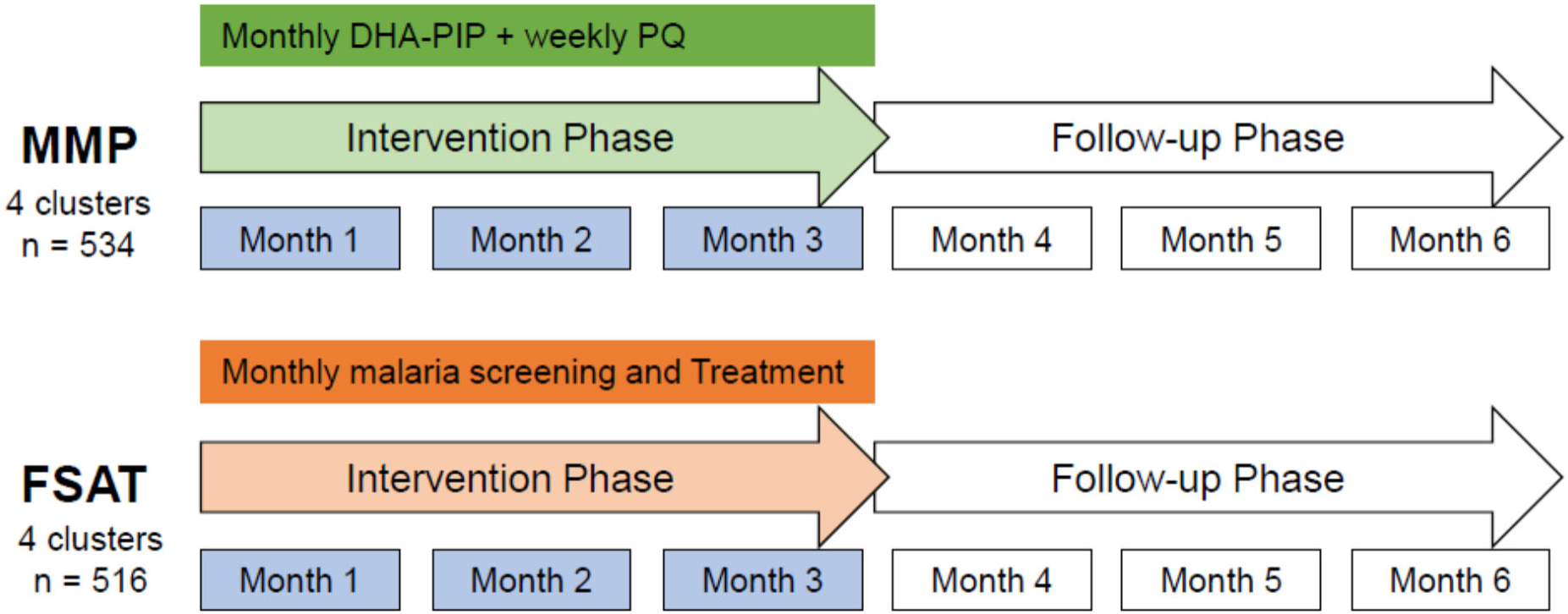
Study design and timeline. All volunteers were screened for malaria with monthly malaria RDT, microscopy and PCR testing. During the three-month intervention phase, volunteers in the MMP treatment arm received regularly scheduled treatment with a full monthly DP treatment course and a weekly dose of 22.5mg primaquine regardless of malaria positivity for a total dose of 270 mg. Volunteers in the FSAT arm only received treatment if found to be malaria-positive by microscopy or PCR. All volunteers in the study were screened monthly for three additional months or when symptomatic with RDT, PCR and microscopy and treated for malaria if positive. Treatment with standard-of-care medications was given if positive by microscopy or PCR within 24-48 hours. Standard doses of PQ were used per national treatment guidelines in the FSAT arm.

### Recruitment

Sites selected were based at military units willing to participate in the study and authorized to do so by their respective Command authorities. Authorization was granted on a collective basis to military personnel at each participating cluster prior to selection. Individuals at each site were enrolled based on willingness to participate, and authorization for military personnel by their supervisor. However, the Chain of Command was not involved in the enrollment process nor permitted to encourage or order soldiers to participate in the study. Independent civilian ombudsman were present during consent and enrollment to minimize undue influence among participants. Study sites were stratified *a priori* into high and low malaria transmission areas based on internal RCAF malaria surveillance data from prior years.

### Study Procedures

Following informed consent or parental consent with assent by minors, all subjects underwent baseline assessments including a history and physical exam, complete blood count, and malaria diagnostic testing. Glucose 6-phosphate dehydrogenase (G6PD)-deficiency screening was assessed using FDA-approved quantitative testing methods (Trinity Biotech, Ireland). Malaria diagnostics included a rapid test (SD BIOLINE Malaria Ag *Pf/Pv* malaria antigen or MALACHECK Ag pLDH/HRP2), microscopy and molecular testing with species-specific PCR using previously described methods [29]. A threshold of <35 PCR replication cycles was used to define positive results for malaria. The study consisted of a three-month intervention period with MMP or FSAT, followed by a three-month follow-up period, during which all volunteers continued monthly screening and treatment.

Malaria treatment was administered according to 2016 Cambodian National Treatment Guidelines (NTG). Children <13 years old were not treated with PQ and had only finger stick blood collections for malaria diagnostics. All volunteers with blood-stage parasitemia by light microscopy were followed to clinical symptom resolution with two negative blood smears at least one week apart. If volunteers were only PCR positive, they were evaluated again the following month to ensure clearance. Recurrent cases of *Pf* were evaluated with molecular genotyping of MSP-1, MSP-2, and GLURP allelic variants to distinguish recrudescence from reinfection [30].

### Study Interventions

Starting at enrollment, volunteers in the MMP group received 3 monthly three-day courses of fixed-combination DP tablets (Doucotexcin, Holley Pharmaceuticals, China) containing 40 mg of DHA and 320mg of piperaquine phosphate (PIP) during the intervention. All MMP volunteers age ≥13 years also received 12 weekly PQ 22.5mg tablets (270 mg total dose) to prevent malaria transmission and *Pv* relapse. MMP participants that were found to be initially positive by PCR at enrollment received their first scheduled course of DP with the prescribed MMP regimen. However, MMP volunteers subsequently found to be parasitemic by PCR or microscopy during follow-up were treated with standard-of-care antimalarials following national treatment guidelines (NTG). Volunteers in the MMP group who received rescue treatment following NTG, were treated with their regularly scheduled MMP regimen with DP on next monthly follow up, and while weekly low dose primaquine was continued on schedule without interruptions.

During the intervention period, all participants in FSAT clusters were screened monthly by RDT, microscopy and PCR and treated following national guidelines if positive by microscopy or PCR. All volunteers in FSAT clusters with parasitemia by PCR or microscopy were treated following the 2016 Cambodia National Treatment Guidelines. *Pf*-positive volunteers and those with mixed-species infections received a full 3-day treatment course of artesunate and mefloquine (ASMQ, Guilin Pharmaceutical Co Ltd, Guilin, China) with a single low 15mg PQ dose (Government Pharmaceutical Organization, Bangkok, Thailand). *Pv*-positive volunteers received a full 3-day treatment course of DP plus 15mg PQ daily for 14 days to prevent relapse. G6PD-deficient *Pv*-positive volunteers received 8 weekly 45 mg doses of PQ. The intervention and follow-up period used the same intervention for FSAT subjects. The follow-up period for the MMP volunteers mirrored the FSAT intervention and follow-up period, reflective of the NTG. All treatment was administered as Directly Observed Therapy (DOT) by RCAF medical personnel on the study team.

Participants were provided with outer garments to wear in forested areas prior to study start. These included civilian clothing or military uniforms as appropriate. Garments were pre-treated with insecticide by the study team prior to distribution using a single application of 40% permethrin Individual Dynamic Absorption kits (NSN 6840-01-345-0237; U.S. Army) to produce treated clothing (ITU) or water to produce sham-treated clothing (sITU). According to the product label, permethrin treatment was designed to withstand 30 washings.

### Statistical Analysis

Detailed description of the statistical plan is outlined in the prior study protocol publication [29]. In brief, the primary study endpoint the absolute risk reduction (ARR) based on the proportion of volunteers in each arm with PCR-corrected absence of parasitemia at the end of 6 months using an Intention-to-Treat analysis. Power analysis to determine the minimum sample size for the study assumed a conservative value of 0.4 for the intra-class correlation coefficient (ICC). It was determined at least 78 individuals were required to achieve 80% power to detect a two-fold reduction in the ARR for MDA over FSAT with an α=0.05. ARR was based on the proportion of malaria-free volunteers at six months [31]. As the number of available clusters was limited for practical reasons, a total of eight clusters with 80 to 140 volunteers per cluster was determined to be sufficient for this pilot study.

Demographic, epidemiological, and laboratory data were summarized at baseline and follow-up. All enrolled volunteers were included in the database for primary endpoint analysis. The safety analysis database included all volunteers who received at least one dose of study drug. T-tests and chi-square tests were used to assess the statistical significance of differences in two means (including log-transformed) or proportions, respectively. Confidence limits (95%) for means, geometric means, and proportions were calculated. A mixed-effects logistic regression model was used to assess the role of treatment group (MMP vs. FSAT), insecticide-treated uniforms (ITU vs. sITU), cluster regional endemicity (low vs. high), and parasitemia at enrollment (positive vs. negative) on the likelihood of a *Pf* or *Pv* infection during the 6 month study period. Random effects were modeled at the cluster level. Time to event data was summarized using Kaplan-Meier plots. The log-rank test was used to assess the statistical significance of differences between the treatment groups.

### Ethics Statement

The trial was approved by the Walter Reed Army Institute of Research Institutional Review Board (WRAIR IRB, Protocol number, WR2211) and the National Ethics Committee for Health Research (NECHR) in Cambodia. The study was registered on Clinicaltrials.gov prior to enrollment (NCT02653898).

## Results

### Enrollment and Study Follow up

Of the 1,114 volunteers residing in the 8 selected clusters who were screened, 1,050 enrolled into the study (**Figure 2**). This corresponded to 94.3% overall coverage in the population studied, ranging from 89-97% coverage within individual clusters. The first subject was enrolled in January 2016 and the last subject follow-up was completed in August 2016. There were 534 volunteers in four clusters assigned to MMP (264 received ITU and 270 sITU) and 516 volunteers in four clusters assigned to FSAT (240 received ITU and 276 sITU). Among enrolled volunteers, 75.8 % (796/1050) were military personnel, 12.8% (134/1051) were non-military adults, and 11.4% (120/1050) were children < 18 years old. All volunteers identified ethnically as Khmer, and 84.3% (885/1050) of the population were male. At six months follow up, 90.1% (485/534) in MMP and 86.0% (443/516) in FSAT groups completed all study follow-up (**Supplemental Table 1**). Overall, only 11% (124/1050) withdrew from the study before completing all procedures, generally due to geographic reassignment or to new duties which precluded continuation of follow up.

**Figure 2.**
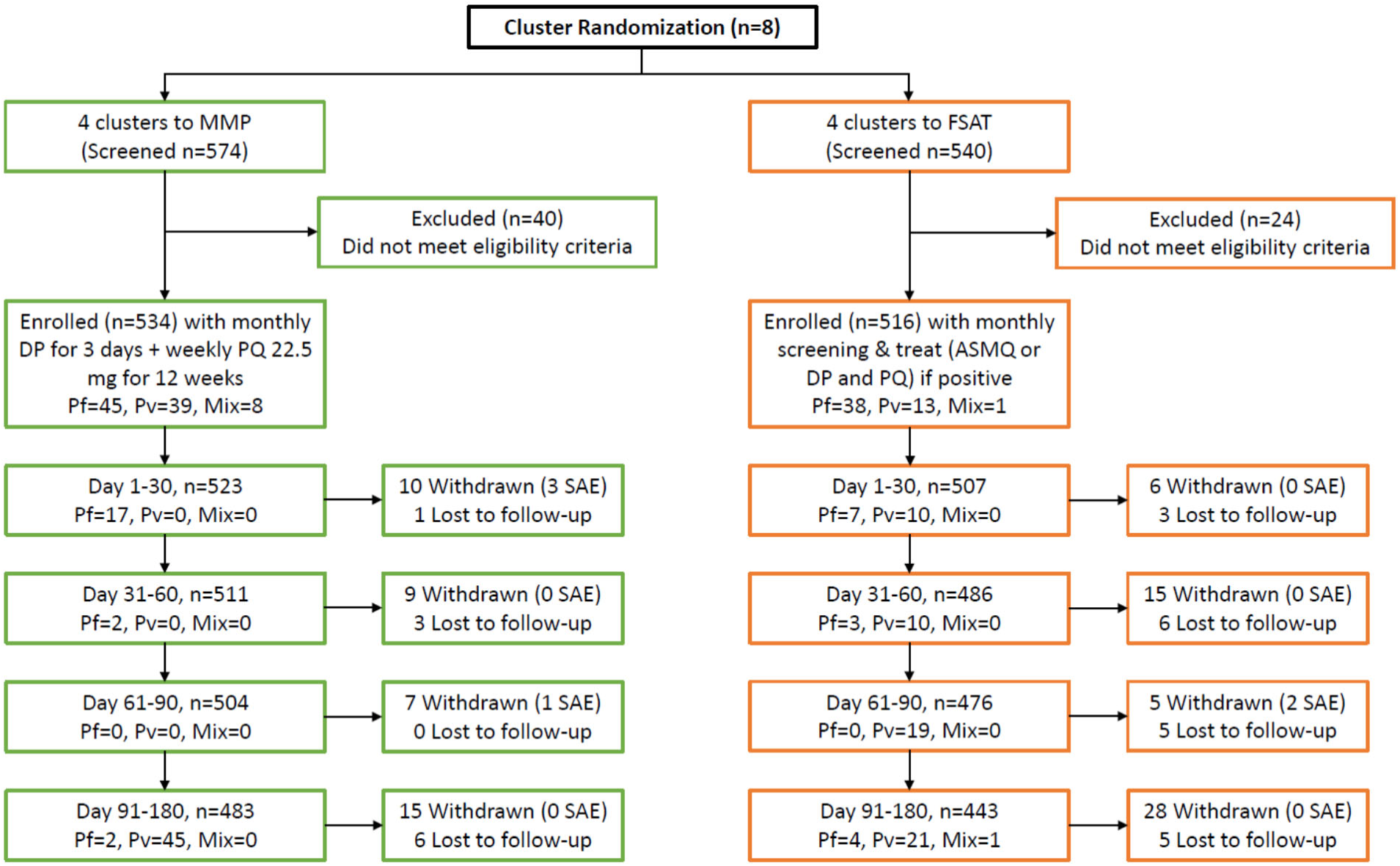
CONSORT Flow Diagram. Volunteers were randomized to one of 8 geographically isolated unit-based clusters. There were a total of 4 clusters for MMP (green) and another 4 clusters for FSAT (orange) treatment interventions. Half of the volunteers assigned to MMP and FSAT were also treated with insecticide treated uniforms (ITUs). Total follow-up was 180 days after enrollment. Most volunteer withdrawals and losses to follow-up were due to movement outside the study areas.

Cluster composition at baseline is shown in **Table 1**. The study was designed to ensure balanced representation of all conditions (treatment, uniform, and endemicity) across the clusters within the study. Baseline Pv prevalence was 8.4% (45/534) in the MMP groups compared to only 2.7% (13/516) in FSAT clusters. Pf prevalence for both arms was comparable at baseline (*Pf* MMP = 62/534 = 9.7%; *Pf* FSAT = 42/516 = 8.1%). PCR-confirmed malaria prevalence at baseline was 15% (145/1050) across all clusters, with *Pf* the predominant species at 8-10%. The MMP clusters had higher baseline malaria prevalence with 92 positive individuals consisting of 45 *Pf* cases (49%), 39 *Pv* cases (42%), and 8 mixed *Pf/Pv* infections (9%) by PCR. Of the 53 PCR-positive cases at baseline in the FSAT clusters, there were 39 *Pf* cases (73%), 13 *Pv* cases (25%) and 1 mixed *Pf/Pv* infection (2%). Withdrawals and losses to follow-up were not evenly distributed among study groups, with a higher proportion withdrawing from the FSAT group (14.5%), compared to the MMP group (9.3%). Differences were nearly 2-fold higher after month 2 with 10.5% lost or withdrawn in the FSAT groups vs. 5.6% in MMP (**Supplemental Table 1**).

### Treatment outcome by malaria species

All groups experienced rapid *Pf* malaria reduction within 30 days of study initiation (Fig 3A and B). At the last follow up on Day 180, less than <1% of the study population had PCR-confirmed *Pf* malaria, with *Pv* being the predominant species at 2-4% (**Figures 2, 3 and 4**). During the study period, based on the detected malaria cases, the predicted unadjusted prevalence of *P. falciparum* was 11,048 episodes/100,000 population and the unadjusted prevalence of *P. vivax* was 15,238 episodes/100,000. From Month 1 to study end, 11.0% (95% CI: 8.0-11.7%) of volunteers experienced *Pv* infection and 15.4% (CI: 11.5-15.7%) of volunteers experienced *Pf* infection at least once.

**Figure 3.**
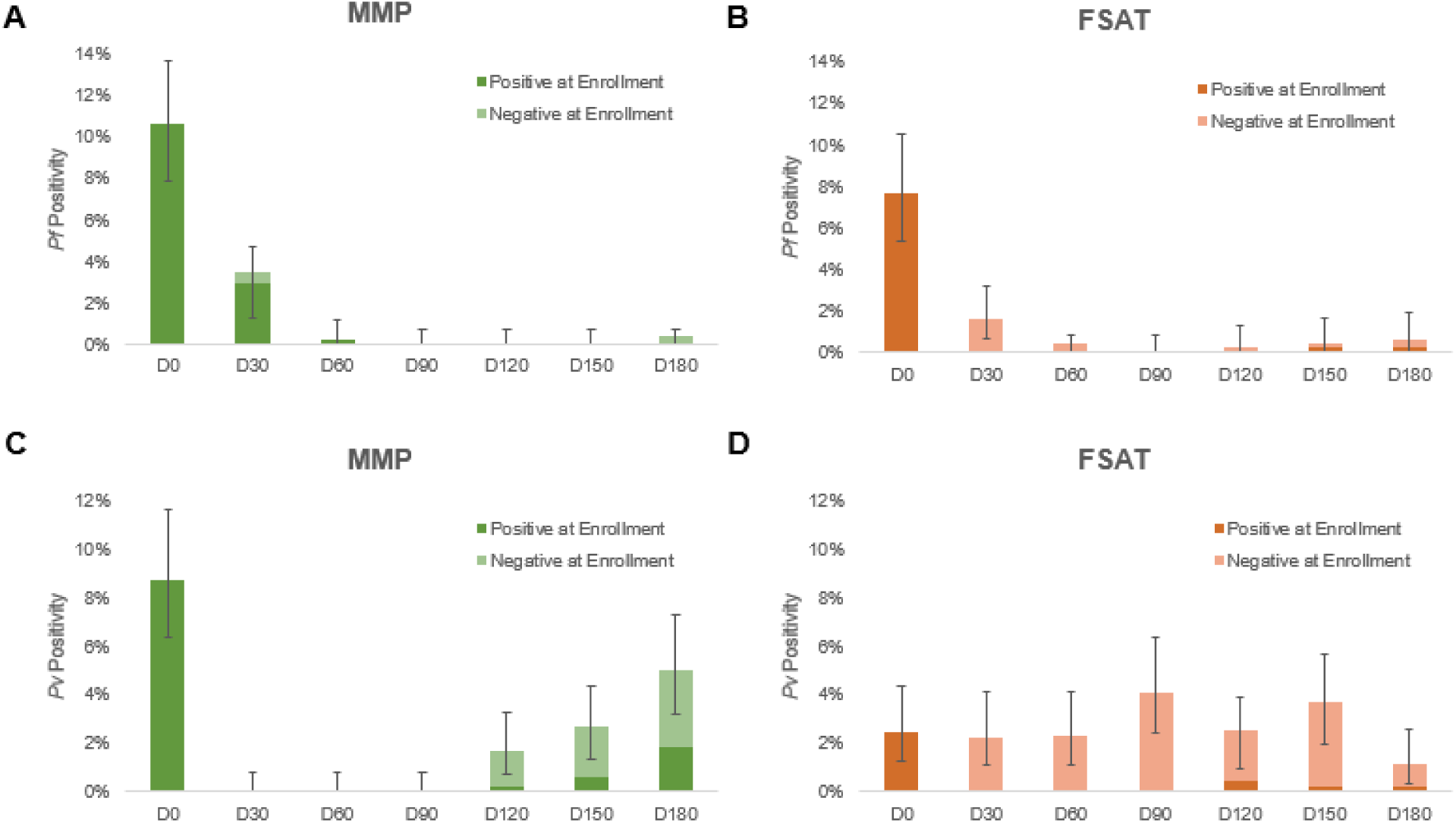
PCR-adjusted malaria prevalence on the monthly follow up in MMP and FSAT. Rapid and sustained reductions in the number of *Pf* malaria cases were observed in MMP (green) and FSAT (orange) (Panel A-B). A high rate of *Pv* malaria was detected for both interventions arms (Panel C-D) despite 270 mg total PQ dose in MMP arm (G6PD normal and G6PD deficient volunteers), over a 12 week period, and conventional doses of PQ in FSAT arm (i.e., 15 mg x 14 days in G6PD normal volunteers and 45 mg weekly x 8 weeks in G6PD deficient volunteers). Results are stratified between subjects that were positive at enrollment and tested positive at a follow-up time point and subjects that were negative at enrollment but tested positive at a follow-up time point. Error bars reflect the 95% confidence interval.

**Figure 4.**
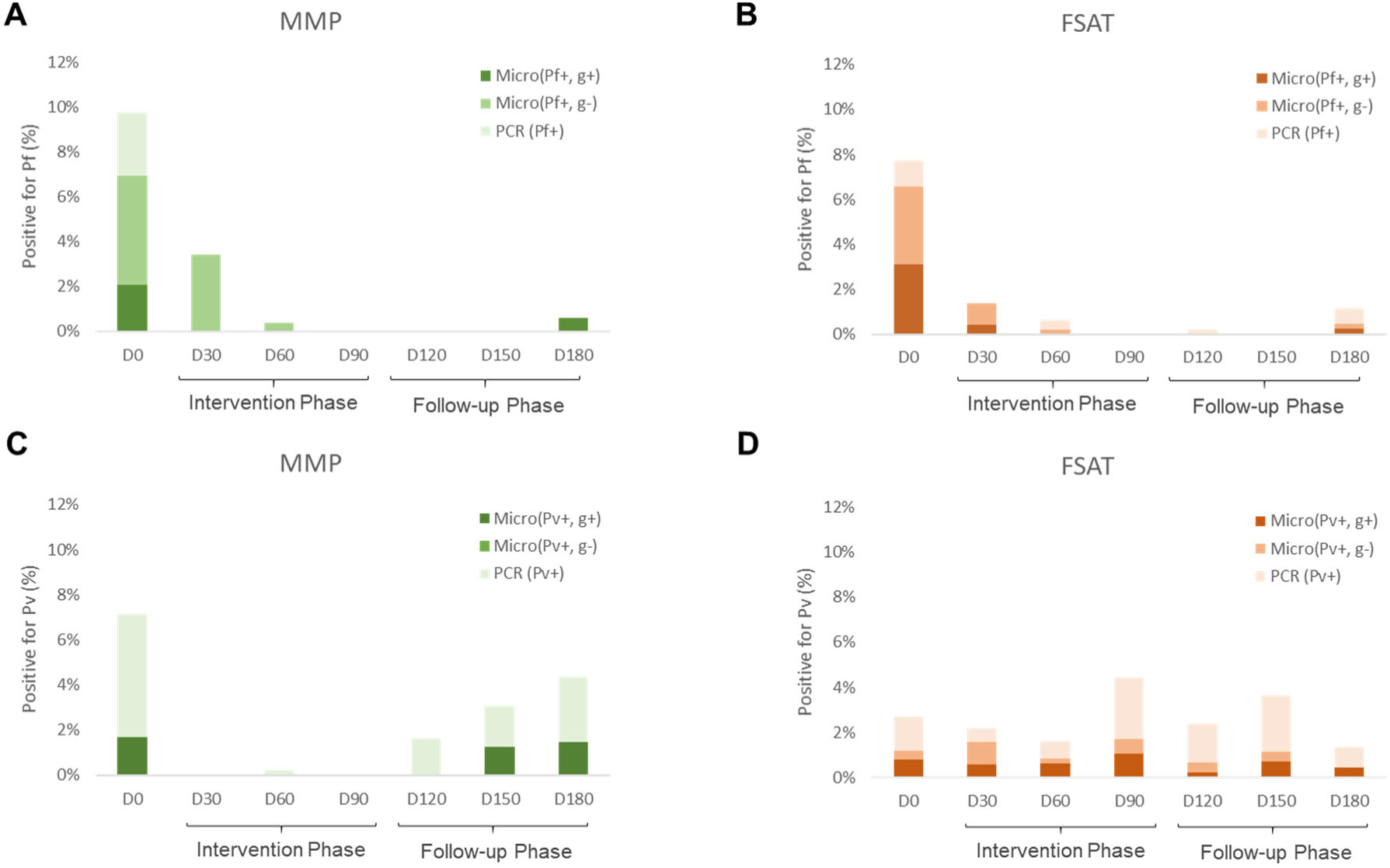
Microscopic gametocytemia by group for *Pf* and *Pv*. Presence (g+) or absence (g-) of gametocytes by microscopy was assessed for each individual at each time point. At least 200 oil immersion fields were examined on a thick blood smear before it was considered negative. Panels **A.** and **B.** compare rates of microscopic gametocytemia between the FSAT and MMP groups for *Pf* gametocytes, while panels **C.** and **D.** compare rates for *Pv* gametocytes. Overall rates of infection by PCR are indicated by superimposed tan (MMP) or salmon (FSAT) colored bars.

*Pf* malaria transmission was interrupted by both MMP and FSAT interventions. PCR-corrected prevalence of *Pf* malaria declined from 9.9% at baseline to 0.4% on month 6 follow up (96% decline) for MMP and from 7.6% to 0.97% (87.1% decline) in FSAT treatment group (**Figure 3, Panel A-B**). In MMP, 16 of 51 (31%) subjects that were positive for *Pf* at enrollment presented with malaria recurrence within their first two months of follow-up, with all 16 genotyped as recrudescences. All recurrent cases of *Pf* in the MMP group were successfully treated with artesunate-mefloquine at the 1-month follow-up visit. In FSAT, where all *Pf*-positive volunteers were treated with artesunate-mefloquine, 2/39 (5%) had PCR-confirmed recurrence within their first month of follow-up, and all cases were confirmed to be new infections. Volunteers assigned to the MMP arm experienced 2 cases of *Pf* in the last 3 months of follow-up, compared to 4 cases of *Pf* in FSAT arm, during the same follow-up period.

In contrast to *Pf* infections, ongoing *Pv* infections were observed in both treatment groups after enrollment during the study period. Despite rapid *Pv* decline in MMP, with no new cases detected during the treatment period (Days 0-90), 45 of 481 (9.3%) MMP subjects tested positive for *Pv* parasitemia after discontinuing prophylaxis within the 90-day follow-up period. In MMP subjects, 29% (13 of 45) of *Pv* cases during the follow-up period were in subjects that were positive for *Pv* at enrollment (**Figure 3, Panel C**). In FSAT, overall *Pv* rates were twice as high as in MMP subjects (cumulative 81 of 444, 18.2%) as in **Figure 3, Panel D**. *Pv* was more evenly distributed in the FSAT groups throughout the study period, with an average rate of 3.0% per month. *Pv* recurrences in individuals that were *Pv* positive at baseline made up a larger percentage of total *Pv* cases in the MMP group compared to the FSAT group (29% vs. 4.9%). The probability of *Pv* recurrence by 6 months if *Pv*-positive at baseline was similar for both groups (30% in MMP and 29% in FSAT). In contrast to MMP, only 4 of 81 *Pv* cases (4.9%) detected during follow-up in FSAT clusters were in *Pv*-positive individuals at enrollment.

The primary endpoint of the study was absolute risk reduction (ARR) for malaria at 6 months. A summary of absolute and relative risk reductions (RRR) are shown in **Table 2**. Overall there was a 68.7% cumulative relative-risk reduction (RRR) for *Pf* malaria in MMP groups at day 90 (0.6% prevalence) compared to FSAT (1.91%) or an absolute risk reduction (ARR) of 1.3%. RRR for *Pf* at day 180 in MMP clusters (1.0%) was 61.5% with an ARR of 1.7% compared to FSAT (2.7%) (Table 2). Overall RRR for *Pv* in MMP groups at day 90 (0% prevalence) was 100% compared to FSAT (9%) or an ARR of 9%. RRR for *Pv* was 47.3% at day 180 in MMP clusters (6.7% prevalence) compared to FSAT (12.6% prevalence) for an ARR of 6%.

**Table 2.**
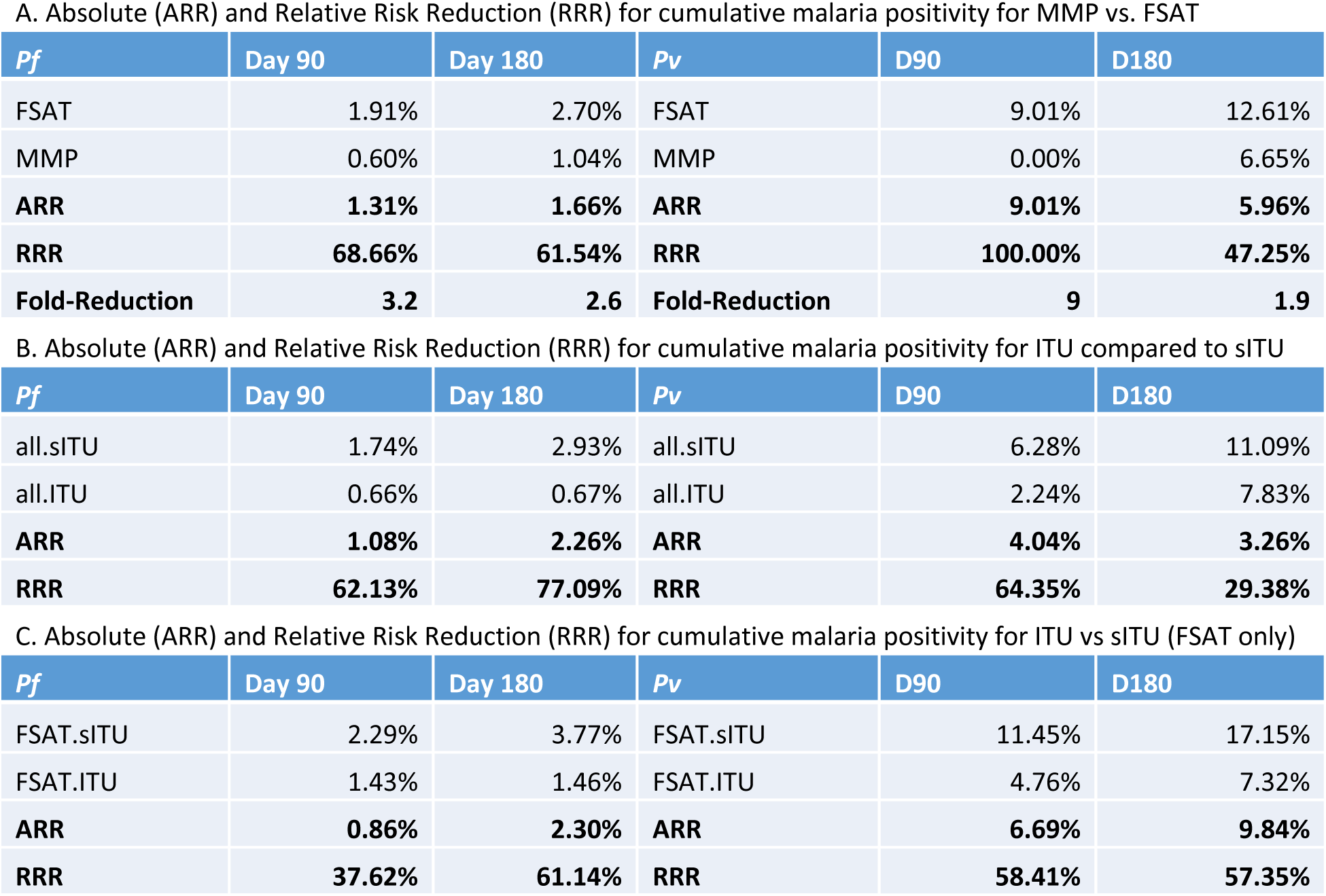
Summary of absolute (ARR) and relative risk reductions (RRR) based on primary intervention (MMP vs FSAT) and uniform treatment (ITU vs sITU). ARR and RRR are calculated based on cumulative PCR-confirmed malaria positivity at months 3 and 6 over the course of the study for A. MMP with respect to FSAT; B. ITU with respect to sITU for both MMP and FSAT; C. ITU compared to sITU for FSAT only.

| *Pf* | Day 90 | Day 180 | *Pv* | D90 | D180 |
| --- | --- | --- | --- | --- | --- |
| FSAT | 1.91% | 2.70% | FSAT | 9.01% | 12.61% |
| MMP | 0.60% | 1.04% | MMP | 0.00% | 6.65% |
| **ARR** | **1.31%** | **1.66%** | **ARR** | **9.01%** | **5.96%** |
| **RRR** | **68.66%** | **61.54%** | **RRR** | **100.00%** | **47.25%** |
| **Fold-Reduction** | **3.2** | **2.6** | **Fold-Reduction** | **9** | **1.9** |

B. Absolute (ARR) and Relative Risk Reduction (RRR) for cumulative malaria positivity for ITU compared to sITU
| *Pf* | Day 90 | Day 180 | *Pv* | D90 | D180 |
| --- | --- | --- | --- | --- | --- |
| all.sITU | 1.74% | 2.93% | all.sITU | 6.28% | 11.09% |
| all.ITU | 0.66% | 0.67% | all.ITU | 2.24% | 7.83% |
| **ARR** | **1.08%** | **2.26%** | **ARR** | **4.04%** | **3.26%** |
| **RRR** | **62.13%** | **77.09%** | **RRR** | **64.35%** | **29.38%** |

**Table 2. Summary of absolute (ARR) and relative risk reductions (RRR) based on primary intervention (MMP vs FSAT) and uniform treatment (ITU vs sITU).**
| *Pf* | Day 90 | Day 180 | *Pv* | D90 | D180 |
| --- | --- | --- | --- | --- | --- |
| FSAT.sITU | 2.29% | 3.77% | FSAT.sITU | 11.45% | 17.15% |
| FSAT.ITU | 1.43% | 1.46% | FSAT.ITU | 4.76% | 7.32% |
| **ARR** | **0.86%** | **2.30%** | **ARR** | **6.69%** | **9.84%** |
| **RRR** | **37.62%** | **61.14%** | **RRR** | **58.41%** | **57.35%** |

Figure 4. shows gametocytes detected by microscopy for *Pf* (panel **A, B**) and *Pv* (panel **C, D**). No gametocytes were seen by microscopy in MMP clusters from months 1-6 for *Pf*, or months 1-4 for *Pv*. While no microscopic *Pf* gametocytes were seen from months 2-6 in the FSAT group, *Pv* gametocytes persisted throughout the observation period at a low level.

### Diagnostic Parasite Detection

Participants were followed with parasitemia assessed monthly by rapid diagnostic test (RDT), microscopy (Micro) and plasmodium species-specific polymerase chain reaction (PCR) for 180 days.

Diagnostic test results are shown in Figure 5 for the MMP and FSAT groups (**5A-B**) and *Pf* and *Pv* for both groups (**5C-D**). Using PCR as the reference standard among 6,784 overall tests, microscopy was highly specific for both *Pf* (99.9%) and *Pv* (100%), but far more sensitive for *Pf* (84.3%) than *Pv* (39.0%). Microscopy was less sensitive than PCR, for both blood stage disease and gametocytes (3E-F). RDT was also highly specific for *Pf* (99.9%) and *Pv* (100%) compared to PCR but had poor sensitivity for *Pf* (69.4%) and virtually no sensitivity for *Pv* (0.5%).

**Figure 5.**
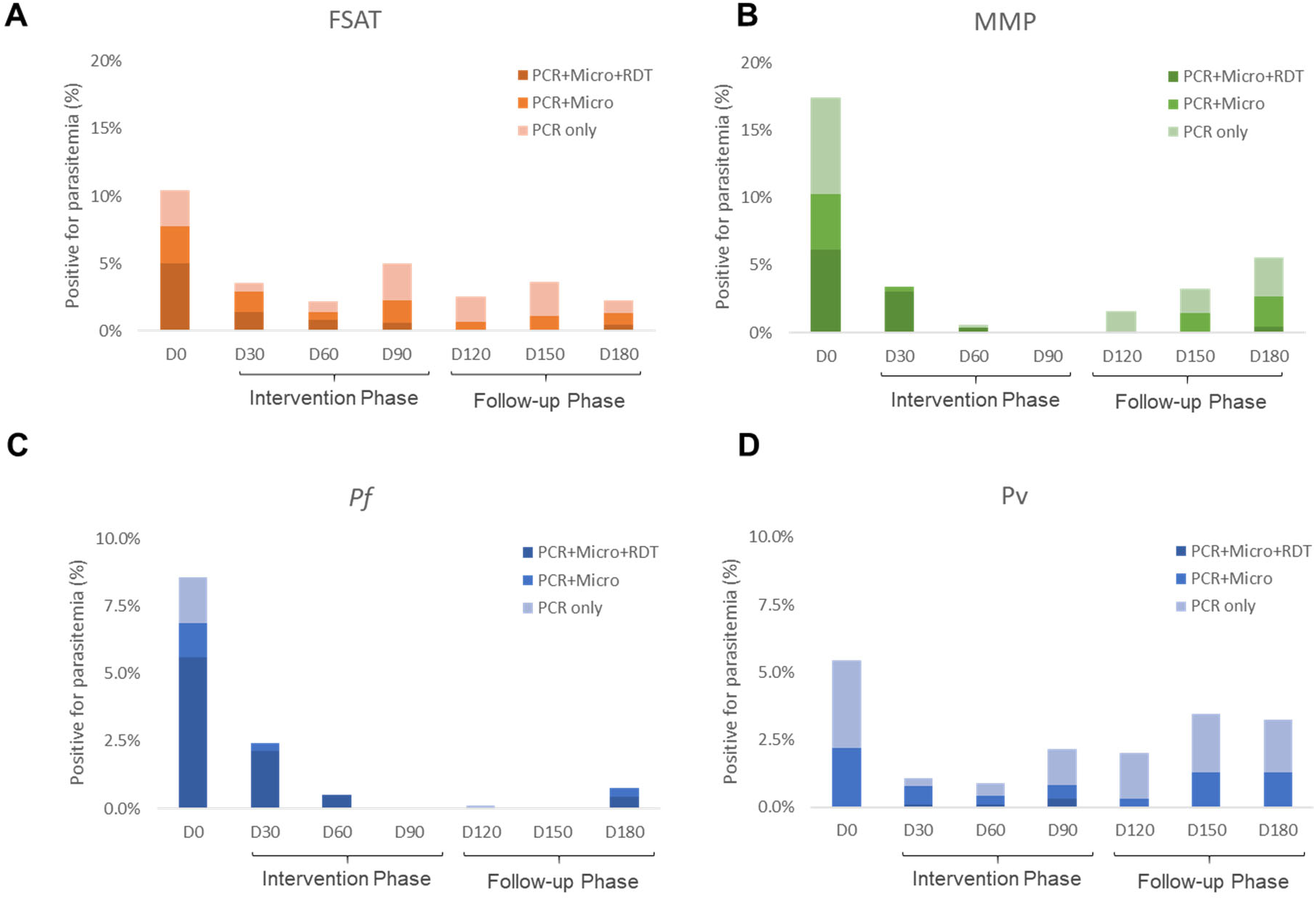
Diagnostic detection of malaria. Prevalence of all malaria species at each monthly follow up is shown based on the three diagnostic methods (RDT, microscopy, and PCR) performed on each individual at each visit. Participants were followed with parasitemia assessed monthly by rapid diagnostic test (RDT), microscopy (Micro) and plasmodium species-specific polymerase chain reaction (PCR) for 180 days. All RDT and Micro-positive cases were also PCR positive. Panels A. and B. compare detection rates between the (A) FSAT and (B) MMP groups. Panels C. and D. compare detection rates for all volunteers for (C) *P. falciparum* and (D) *P. vivax*.

### Effect of treated uniforms and endemicity on treatment outcome

Mixed-effects logistic regression was used to assess effects of uniform treatment and cluster endemicity on treatment outcome. Endemicity and uniform treatments are expected to affect the rate of acquiring *new* infections by affecting malaria vector exposure rates, but not rates of relapse or drug treatment failure. As such, analysis included only subjects who were negative at enrollment for *Pf* or *Pv*, respectively, to assess the role of treatment and endemicity without confounding from treatment failure or relapse. Overall, for *Pf* infections, FSAT had higher cumulative *Pf* positivity at 6 months than MMP regardless of endemicity or uniform conditions (p < 0.01) (Figure 6A). Although uniform treatment had no statistically significant effect of on cumulative *Pf* positivity, the total number of *Pf* cases in the study after Day 30 was low (16 total cases when excluding subjects positive at enrollment). There were no cases of *Pf* during follow-up in MMP subjects with treated uniforms. Cumulative *Pf* positivity in FSAT for those with sham-treated uniforms (3.9%) was approximately double that of those with permethrin-treated uniforms (1.9%; RRR 61%; ARR 2.3%).

**Figure 6.**
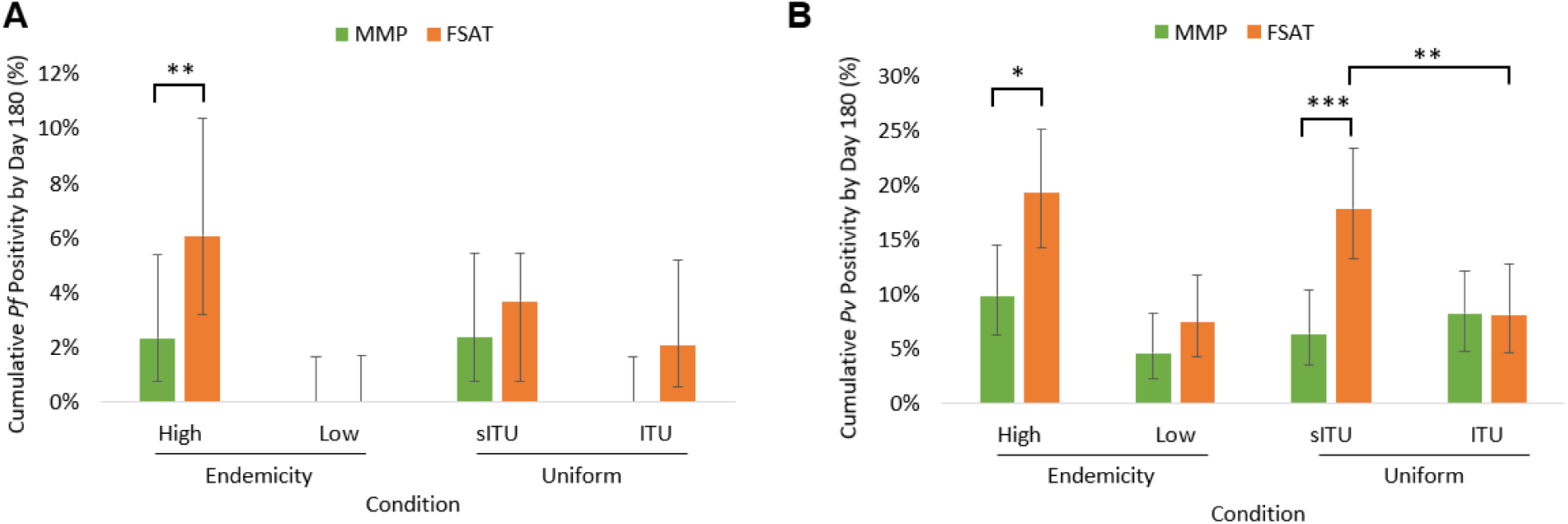
Cumulative *Pf* and *Pv* positivity with respect to drug intervention, uniform treatment, and endemicity. Cumulative positivity from Day 0 to 180 is shown for *Pf* (A) and *Pv* (B); MMP (green bars) and FSAT (orange bars) treatment conditions, stratified by cluster endemicity (high vs. low) and uniform (sITU vs. ITU). 95% confidence interval error bars and statistical significance (* p < 0.05, ** p < 0.01, *** p < 0.001) are shown.

There were significant effects of endemicity and uniform treatment on cumulative Pv positivity (Figure 6B). Cumulative *Pv* positivity was approximately double in clusters with high endemicity compared to clusters with low endemicity. Stratifying for endemicity, FSAT subjects had significantly higher *Pv* positivity than MMP subjects in high endemicity clusters (p < 0.05) but not low endemicity clusters. Stratifying by uniform type, FSAT subjects with sham-treated uniforms had almost 3-fold higher cumulative *Pv* positivity than MMP subjects with sham-treated uniforms (p < 0.001). However, there was no difference in *Pv* positivity between MMP and FSAT subjects wearing treated uniforms. FSAT subjects wearing sham-treated uniforms had almost 2.5-fold higher *Pv* positivity at 3 (6.3%) and 6 months (17%) than FSAT subjects wearing treated uniforms (2.2% and 7.3% respectively; RRR=57.4% and ARR=9.8% at 6 months; p < 0.01). See **Supplemental Table 2** for summary logistic regression results.

### Malaria infection rate over time

Survival analysis was performed to evaluate rates of acquiring *Pf* and *Pv* infections over the course of the study. To ensure that the analysis was focused on new infections and not confounded by relapse or treatment failure, malaria positive subjects at enrollment were excluded. Time-to-event analysis was based on time to acquire the first documented *Pf* or *Pv* infection for each subject. The analysis was stratified by endemicity, and included only high endemicity clusters for *Pf* as no new *Pf* infections were observed in low endemicity clusters. *Pv* survival curves were stratified by uniform assignment (sITU vs ITU) after logistic regression analysis showed endemicity and uniform were significantly associated with cumulative *Pv* positivity.

Survival analysis showed significantly fewer *Pf* infections in the MMP than FSAT groups (Figure 7A, p<0.05), with no significant differences seen based on uniform treatment (not shown). For the *Pv* survival curve in the high endemicity cluster, MMP treatment had significantly lower infection rates than FSAT treatment (p<10^-5^). Treated uniforms showed significantly lower infection rates than untreated uniforms (p<0.05), with an interaction observed between treatment and uniform type (p<0.05). Overall, in the high endemicity conditions, the FSAT+sITU condition performed significantly worse than the other three conditions (p < 10^-7^). The *Pv* survival curve in low endemicity clusters showed no statistically significant difference in infection rates for treatment or uniform type. However, there was a trend with higher infection rates for the FSAT+sITU condition than the other three. It is important to note, in the *Pv* survival curves, that while FSAT infection rates remained steady, the MMP infection rate was zero until D90, after which it increased to a level comparable to FSAT.

**Figure 7.**
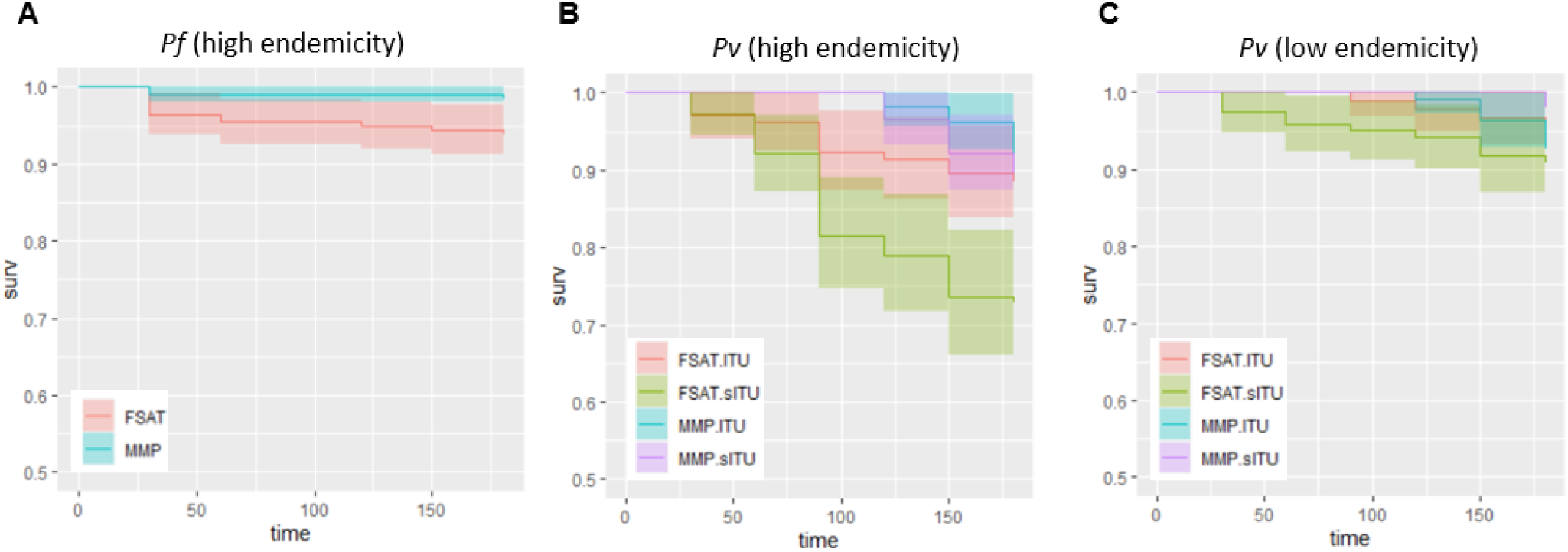
Survival curve to first *Pf* or *Pv* infection stratified by treatment. Survival curves are shown for *Pf* infection for FSAT (red) and MMP (blue) treatment conditions for in high endemicity clusters (A). Survival curves for Pv infection in high and low endemicity clusters (B and C) are shown for FSAT+ITU (red), FSAT+sITU (green), MMP+ITU (blue), and MMP+sITU (purple) conditions. 95% confidence intervals are shown as shaded colors corresponding to each condition. There was only 1 low endemicity *Pf* case which is not graphically displayed.

### Safety

Of 1,050 volunteers enrolled, there were 1,237 adverse events (AEs) reported over the 6-month study or 1.2 AEs per volunteer. The vast majority of AEs (88.0%) were classified as unlikely or not related to study interventions. The most frequent AE was upper respiratory infection. Most AEs were mild to moderate in severity, and all resolved prior to study discharge. Study drugs were well tolerated, and there were no AEs reported from ITU wear. There were 27 SAEs recorded during the study period. All were unrelated to the study interventions except for three that were classified as ‘unlikely related’ and 1 ‘possibly’ related (new onset migraine headache which resulted in inpatient treatment). All volunteers with SAEs were followed until recovery or until their condition has stabilized. One volunteer was diagnosed with breast cancer and was withdrawn from the study while receiving treatment. One volunteer was withdrawn due to intolerance to DHA-PIP in MMP arm (vomiting). One volunteer died from causes unrelated to study interventions with causality assessed by the research monitor, an independent review, and the ethics review board.

While there have been reports of clinically significant hemolysis observed following PQ treatment even in G6PD normal individuals [32], no significant hemolysis occurred in G6PD-deficient volunteers taking primaquine during the study. All volunteers with moderate to severe G6PD deficiency tolerated the modified 12 weekly doses of PQ without clinically significant hemolysis. There were 160 volunteers (15%) with G6PD-deficiency identified at screening. Of these, 97 (9.2%) took primaquine during the study, but only 14 (1.3%) had a hemoglobin decline >10% at day 3 (**Table 1**). No volunteers had a hemoglobin drop >25% from baseline or required stopping therapy.

## Discussion

At the time the study was designed in 2012, two alternative approaches to malaria elimination were debated in Cambodia. The first, often termed Mass Drug Administration or MDA was thought to be the more effective approach. MDA offers the promise of lower costs, simplified logistics and improved outcomes from sustained pressure on the parasite. However, perceptions surrounding compulsory drug treatment for apparently healthy individuals with unclear risks with low malaria transmission in the GMS raised ethical concerns. A large number of MDA studies have been conducted over the past 70 years with tremendous variation in disease settings and approaches employed. A recent systematic review highlighted difficulties comparing studies of varying designs, often complicated by a lack of well-defined objectives [15]. Despite documented logistical challenges to implementation and lower effectiveness [20], a reactive ‘screen and treat’ approach (which we termed Focused Screening and Treatment or FSAT) was thought more ethically palatable with only confirmed infections given drug treatment. Compared to prior studies that largely relied on less sensitive RDTs or microscopy, treatment in FSAT group in this study was based on PCR results affording greater number of subjects getting treatment.

We sought to reconcile and recast these opposing views to malaria elimination, noting that human malaria often survives in the host undetected in latent, sub-clinical and sub-microscopic forms. Further, in mixed-species transmission settings such as this one, treatment with blood stage antimalarials may not interrupt disease transmissibility, and may in fact potentiate it [33]. Our use of the term monthly malaria prophylaxis (MMP) was deliberate and intended to convey the full therapeutic intent. The MMP addresses the four facets of affective treatment and malaria prevention, namely it (1) targets blood stage infections; (2) reduces transmission risk posed by untreated infection; (3) lowers potential relapse from Pv if treatment is co-administered with primaquine; and (4) lowers the risk of reinfection by reduction of malaria burden. The treatments employed were intended to reduce and ultimately interrupt malaria transmission with limited need for micromanagement required with a screening and treatment approach. Namely frequent and repeated clinical evaluation, safety and molecular malaria diagnostic tests, and therapeutic interventions. Insecticide-treated clothing was assessed for added benefit against day-biting mosquitoes in endemic forested areas.

In the MMP regimen, the four therapeutic objectives above were achieved using dihydroartemisin-piperaquine to treat acute parasitemia and provide suppressive chemoprophylaxis (Objectives 1 and 4), and low-dose weekly primaquine to reduce transmission risk, eliminate or suppress latent Pv infection (Objective 3), and provide causal liver-stage chemoprophylaxis (Objectives 2, 3, and 4). MMP groups reached zero PCR-detectable malaria parasitemia after 3 months of treatment. There was no microscopic gametocytemia detected between Months 2-6 or blood-stage *Pv* by PCR between Months 2-4. There were relative risk reductions (RRR) of 60-70% in cumulative *Pf* infection in the MMP group compared to FSAT over the course of the study, with a 1.6% absolute risk reduction (ARR). The RRR of MMP relative to FSAT was 100% for *Pv* after the three-month intervention period, and 47% through 6 months of follow-up with an ARR of 6%. Uptake of therapy was excellent, well-tolerated, and coverage was high in this voluntary study. The primary drawback for MMP use was high baseline *Pf* resistance to DP in Cambodia. The ∼30% clinical failures required rescue therapy with A+M at Month 1, though all were cured and remained *Pf* negative thereafter. Future studies could investigate the use of other drug combinations that are more effective for MDR *Pf* strains in Cambodia which would eliminate the need for rescue treatments and monitoring for recrudescence.

In retrospect, the MMP results could have been achieved in this endemic community with less frequent monitoring than a reactive screen-and-treat approach. MMP findings were in contrast to those among the FSAT groups, particularly for *Pv*. The *Pv* parasitemia in MMP remained endemic at a rate of 2-4% throughout the study. This was despite a 4-fold lower baseline *Pv* infection rate in FSAT clusters (∼2%) compared to MMP (>8%). During the follow-up period when MMP and FSAT groups had the same screening/treatment regimen, malaria re-emerged in the MMP group, with *Pv* parasitemia detected within one month and *Pf* within three months after the intervention period. Although the study was not powered to assess the effectiveness of insecticide-treated clothing, there was a statistically significant benefit against *Pv* infection in the FSAT but not MMP groups. Uniform treatment may provide additional protection from malaria absent chemoprophylaxis as part of a screen-and-treat approach.

PCR was the definitive diagnostic method in the study, given high sensitivity for both *Pf* and *Pv* species compared to microscopy and RDTs. Microscopy had reasonable sensitivity for *Pf* (84.3%), but poor sensitivity for *Pv* (39%). RDT had poor sensitivity for *Pf* (69.4%) and was almost completely unable to detect *Pv* (0.5%), suggesting little diagnostic utility in elimination. While the PCR method used was implementable from the reference lab supporting the field sites, the technical requirements and cost may render this approach challenging in some settings. Microscopy was able to detect gametocytes unlike the other two methods, providing additional insights into transmission potential. However, gametocyte detection is more labor intensive, requiring longer microscopy reads covering more fields.

A recent systematic review identified at least 10 randomized trials evaluating malaria MDA in high and low incidence settings [15]. In low-transmission settings using various regimens, both *Pf* and *Pv* burden were substantially reduced within 1-3 months. A prior systematic review of 32 studies from 2013 found that MDA led to sustained malaria reductions only in geographically isolated communities [17] [32]. The key lesson from our study, and those preceding it, is the need for sustained intervention beyond three months, recognizing the difficulties comparing interventions from varied populations, risks and settings [34]. Although impact is often relatively short-lived, pharmaceutically driven approaches to malaria elimination appear to be more effective in low intensity transmission settings such as Cambodia (1-10% prevalence). The most challenging elements to account for in any MDA study are movements in and out of clusters. The clusters on the Cambodian-Thai border in our study were not truly geographically isolated from malaria risk areas which tend to be occupational rather than residential. Despite these challenges, the present study sustained parasite-free intervals in MMP clusters longer than most prior efforts with 5 months before PCR-confirmed *Pf* reappeared in the study areas. Both approaches were safe with the MMP approach superior to FSAT for parasite reductions, potential to reduce resource requirements and scalability.

Cambodia has long been among the highest risk countries for antimalarial drug resistance for both *Pf* [35] and *Pv* [36]. Recent high rates of drug failure and tolerability considerations limited possible elimination regimens [4, 37]. Given a lack of options for treatment when the study was implemented, a monthly 3-day DP course was chosen as preferred chemoprophylaxis in the MMP clusters despite clinical DP failures widely reported in the study area three years before study start [38]. At the time of the study, Cambodia had recently replaced DP with A+M as first line *Pf* agent with a single 15mg dose of primaquine to prevent transmission. The recent drug policy change was based on inverse resistance patterns observed between DP and A+M in Northern Cambodia [35]. In line with these findings, there was a 32% initial DP failure rate in the MMP group in the first month, compared to no A+M regimen failures in the FSAT group, and only 3 A+M recrudescences over 6 months. However, all treatment failures in the MMP group were successfully cured using A+M rescue therapy on month 1 follow-up. While a ‘failing regimen’ ultimately managed to suppress *Pf* parasitemia there was a need to surveil for recrudescences. Despite initial DP treatment failures, from an elimination standpoint DP+PQ treatment in the MMP arm significantly outperformed FSAT using A+M treatment (RRR 60-70% for MMP compared to FSAT for cumulative *Pf* during the study period). Our study was not alone in selecting DP as a monthly blood stage elimination agent. The closest recent comparable study used monthly courses of DP with single low-dose PQ to eliminate *Pf* in Vietnam, Cambodia, Laos and Myanmar [39]. The study was larger (∼8,000 volunteers in 16 clusters) but coverage and acceptance were lower with fewer than half of those treated completing 3 months of therapy. Despite the limitations, the approach worked with *Pf* suppression persisting as long as one year in lower transmission areas. We did not find any significant drug resistance at the study sites and observed that 9 of 10 cases of highly resistant *PfPailin* strain were cured with initial therapy. To date there is limited evidence of a sustained MDA or prophylaxis approach generating significant resistance.

*Pv* will likely be the greatest long-term obstacle to malaria elimination in Cambodia and SE Asia [40]. *Pv* constituted 57% of overall malaria and 88% of malaria detected after initial enrollment in the present study. Blood-stage *Pv* prevalence at baseline was comparable to *Pf*, and previously reported relapse rates in the study area were as high as 70-80% in 2010 without primaquine radical cure [2]. Here, MMP effectively suppressed or eliminated blood-stage *Pv* during the three-month intervention period with zero detected cases of *Pv* through Month 3, despite higher baseline Pv prevalence in MMP clusters (8.4% compared to 2.7% in FSAT clusters). By contrast, monthly blood-stage *Pv* rates in the FSAT group remained largely unchanged between 2-4% throughout the study period. There are few studies of *Pv* anti-relapse therapy in elimination settings [41]. Interpretation is confounded by the complexity of distinguishing relapses, recrudescences, and reinfections [42]. In this study, it was impossible to determine whether MMP was effective against *Pv* because it cleared hypnozoites (PQ), interrupted transmission (PQ), reduced reinfection (DP or PQ), or simply suppressed apparent blood-stage relapses (DP). The results based on the timing of infection suggest that MMP *Pv* recurrences were more likely to be the result of relapses, while FSAT recurrences are more likely to be the result of reinfections (Supplemental Figure 2). The positive effect of ITU on *Pv* in FSAT supports the notion that ITU might have helped interrupt *Pv* transmission in those settings. Although a higher percentage of MMP cases were recurrences, there was an overall lower rate of *Pv* in the MMP group. The difference in interval between the two regimens may be due to a combination of factors – a higher reinfection rate of new *Pv* infections in FSAT, suppression of detectable relapse during the intervention period in MMP, or both. However, it is clear that the FSAT approach had little effect on *Pv* rates despite prompt administration of primaquine radical cure following detection. This has implication for traditional malaria control programs, where even if radical cure treatment is administered by DOT, it may be difficult to reduce *Pv* malaria in an expeditious manner. Interpretation may be confounded by successive relapses of unrecognized subclinical disease, indicating a higher proportion of the population affected at baseline than detectable with current diagnostics.

A 22.5mg weekly primaquine dose was previously found to be effective in eliminating latent Pv in immigrants to Australia, and was the basis of the MMP regimen selected here [43]. *Pv* at enrollment was the single greatest predictor of *Pv* positivity during the study using logistic regression, suggesting that relapse played a major role in *Pv* infection rate. In both MMP and FSAT groups, approximately 30% of individuals positive for *Pv* at enrollment had a *Pv* recurrence during the study. This compared to a cumulative *Pv* rate of 7.4% in MMP and 15.6% in FSAT if *Pv* negative at enrollment. These findings suggest that regardless of the different PQ regimens used in MMP and FSAT, the cumulative 270 mg dose of PQ employed here [43] may not be completely effective at preventing relapse. This is in line with a recent study that found that PQ radical cure may only be 60-70% effective at preventing recurrences in this setting [44]. Relapse efficacy may also be affected by a high proportion of intermediate metabolizer genotypes for Cytochrome P450 2D6 polymorphisms [45]. Normal CYP2D6 activity is thought to be required for primaquine to be effective, though clinically significant reductions in primaquine effectiveness have not been conclusively demonstrated in Cambodia.

Malaria elimination efforts are unlikely to be successful without suitable attention paid to the infectious gametocyte. Treatment of blood stage parasites can stimulate gametocytemia and increase transmission risk [33]. Few agents are capable of effectively treating stage V gametocytes in the human host to block mosquito transmission. While artemisinins are thought to have some antigametocidal activity, PQ is the most effective drug for this purpose [46]. A single 45mg dose of PQ administered with blood-stage therapy was a highly effective transmission-blocking agent at similar geographical areas as the present elimination study [47], eliminating circulating *Pf* gametocytes. It also prevented transmission from patient blood samples to laboratory reared mosquitoes compared to no primaquine therapy. A lower single 15mg dose is now widely recommended for transmission-blocking in combination with a blood stage agent. While the MMP and FSAT groups were treated with PQ, the MMP group received a weekly dose of PQ well in excess of the recommended transmission-blocking dose. The FSAT group received a single transmission-blocking dose only if confirmed *Pf* positive or a radical curative dose only if *Pv* positive, leaving subclinical gametocytemia untreated. In the MMP group, no microscopic *Pv* gametocytes were observed from Months 1-4 and no *Pf* gametocytes from Months 1 to 6. In the FSAT group, while no *Pf* gametocytes were observed from Months 2-6, *Pv* gametocytes were seen in 50-67% of FSAT *Pv* infections throughout the entire 6-month observation period, suggesting onging *Pv* transmission throughout the study. This may help explain why the addition of ITU was shown to be beneficial in FSAT where transmission was never interrupted with the drug therapies alone. While gametocytemia was not measured by PCR in the present study, microscopic gametocytemia is thought to have substantially higher transmission risk than sub-microscopic gametocytemia [48]. Unfortunately, assessing community-level impact of transmission blocking agents is extremely challenging [46]. Further, though clusters were geographically isolated by domicile, they were not necessarily isolated in the higher-risk occupational areas. In Cambodia, risks are largely occupational – forested areas patrolled by military personnel in this case [49]. Individuals may have been exposed to infectious bites outside their dwelling clusters, and may have transmitted disease there as well. As such we can only speculate on the relative contribution of transmission blocking effects to the results.

One of the key features distinguishing the study from prior attempts at MDA was our intent to determine effectiveness of chemoprophylaxis in the military in the MMP group as a potential elimination approach, in combination with vector interventions (ITUs). This MDA approach is often considered to be ineffective given risks for adverse events and drug resistance pressure on the parasite [15]. However, some have argued for targeted chemoprophylaxis in forested malaria-endemic areas [49]. The results here largely support that recommendation, suggesting that safe, effective chemoprophylaxis including an 8-aminoquinoline is warranted in this low-transmission, high antimalarial drug resistance setting. A combination of monthly 3-day treatment courses of DP and weekly low-dose PQ safely led to a Relative Risk Reduction of 100% over three months and 47% over six months. While it is impossible to assess the individual contribution of DP and PQ in this study, inferences can be made based on known mechanisms of action. The primary blood-stage post-treatment prophylactic effect was likely due to piperaquine, despite substantial documentation of both clinical artemisinin and piperaquine resistance in this area [4]. While the DP combination initially failed to treat blood-stage disease in >30% of subjects, it appeared sufficient to suppress submicroscopic blood-stage parasitemia during the remaining intervention period. PQ also has ‘causal’ liver-stage antimalarial activity against *Pf* and *Pv*, and there may have been some synergy between DP and PQ in suppressing blood and liver-stage parasitemia [49]. It is impossible to distinguish the potential contributions or synergy of primaquine given the combination approach employed. Others have found similar success with prophylaxis using more limited regimens. A comparison of a monthly treatment course of artemether-lumefantrine showed significant risk reductions compared to a multivitamin in forest-dwellers in Cambodia [50]. The large number of variables in the study obviated establishing a control group for each of the interventions employed.

The study demonstrated the safety and tolerability of the pharmacologic approach to elimination. High compliance was likely achieved through the use of DOT administration. DP is known to be safe and well-tolerated with the exception of elevated risk of QT-interval prolongation when given in a compressed 2-day course [54]. The primary safety concern in the study was use of PQ in G6PD-deficient volunteers. We were the first group to employ the 22.5mg/week PQ dose regimen in an endemic area as an elimination tool to prevent *Pv* relapse. The 22.5mg weekly dose chosen for the MMP group was based on a previously safe, effective anti-relapse dose employed in Asian migrants to Australia [43]. PQ likely provided anti-gametocyte activity and at least some degree of ‘causal’ liver-stage prophylaxis. The FSAT groups used widely recommended PQ doses demonstrated to be safe in G6PD-deficiency for *Pf* transmission blocking and/or *Pv* anti-relapse. In the MMP group, 16% of volunteers were G6PD deficient, comparable to prior observations [55]. Only 15% of subjects had hemoglobin declines of >10% at day 3, none had declines of >25%, and none had to stop PQ treatment based on the pre-specified individual halting rule of Grade 3 hemolysis. This is in line with expectations that even those with relatively severe G6PD deficiency are able to tolerate PQ in low doses [56] and the modified weekly regiment studied.

Vector-based interventions have long been a cornerstone of malaria control and containment. Insecticide-treated bed nets (ITN) have historically been responsible for large reductions in malaria in sub-Saharan Africa. However, ITNs are less effective in Southeast Asia with predominantly day-biting Anopheles species who breed in forested areas, and can be exophagic [49, 57]. Insecticide-treated clothing, typically using wash-resistant permethrin formulations, is often employed by military forces to reduce vector-borne disease risk. The vast majority of vector-based interventions are tested based on their efficacy against vector biting behavior. The study is also among the few to subject a vector-based intervention, particularly insecticide-treated uniforms and clothing to the rigor of a clinical trial of the disease of interest. Only a handful of studies to date have assessed disease outcomes [58], despite emerging concerns for malaria vector permethrin resistance in Africa [27]. Addition of insecticide-treated uniforms (ITU) reduced malaria prevalence when paired with the FSAT but not MMP approaches, suggesting this may be a good options for malaria elimination in select groups. Although difficult with multiple interventions simultaneously employed to definitively distinguish relative contributions to outcomes, the results support ITU use in regions with day-biting vectors. While clothing and uniforms were treated by the study team for this exercise, individual permethrin clothing treatment is relatively easy to perform. ITU use resulted in significantly lower *Pv* positivity rates (p < 0.01) with RRR of ∼60% for *Pv* compared to sITU. A similar effect size was seen for *Pf* with RRR of 40-60%. The latter difference was not statistically significant given the small number of *Pf* infections in the study. It is interesting to note that the observed effect size was consistent for both *Pf* and *Pv*. This was expected as the two species share the same vector and should be equally affected by vector control measures. No statistically significant difference was found in *Pf* and *Pv* infection rates for ITUs vs. sITUs in the MMP group. It is possible that chemoprophylactic effects of the MMP regimen may have masked added beneficial effects of uniform treatment.

This was the first study of a malaria elimination effort in the GMS carried out by and for a military population. The present study demonstrates the effectiveness of the approach described in our prior concept paper on malaria elimination by militaries in the GMS [28]. No elimination scheme can work without a well-trained team able to carry out the complex set of clinical and laboratory activities required. The joint team employed on this study included a large number of Royal Cambodian Armed Forces medical and logistics personnel. The study created a basic Unit of Action for malaria elimination easily replicated elsewhere. The Unit of Action included study physicians, nurses, lab technicians, data analysts and a large cadre of soldier-medics training and working alongside the multinational AFRIMS Malaria team. Achieving and sustaining the required operational tempo required for universal quantitative G6PD testing, frequent diagnostic PCR testing and DOT would not have been possible without significant RCAF support. The study demonstrated that elimination is not only possible within GMS militaries but can be achieved by GMS militaries supporting national efforts. Despite the achievement, those considering the approach must be mindful of the military ethical considerations, and implement appropriate safeguards such as the use of non-military personnel as ombudsmen during the consent process. Consent whether in the context of research or public health implementation, is essential to establish and maintain trust.

There were several limitations to the study. First, despite randomization, the multiplicity of simultaneous interventions makes it difficult to quantify relative contributions of individual interventions. This was a deliberate choice. The goal of the study was to combine what were thought to be the most regionally appropriate, safe, cost-effective elimination strategies available at the time rather than precisely delineate individual contributions of each intervention. Second, assessment of potential reinfection for *Pv* was challenging, given the difficulties in distinguishing relapse, reinfection and recrudescence. Third, we could not control for the effects of in- and out-migration. The former may have introduced additional disease burden to clusters while the latter caused losses to follow-up from occupational reassignment. Fourth, secular declines in parasite prevalence from either seasonal changes or ongoing control efforts in the region were also impossible to quantify. One or more no-intervention clusters may have helped control for this, but were not undertaken for both ethical and practical considerations. To our knowledge, study sites selected did not undergo other control measures beyond those of the study during the observation period. Also, the study concluded during a time of the year generally considered to be a higher malaria transmission period. The enrollment began during low malaria transmission (January) and concluded during the rainy season (June) when malaria transmission in Cambodia typically increases [59]. Seasonality would likely have contributed to a secular rise, rather than fall, in infection rates over the course of the study if the control measures were ineffective. Finally, selection bias may have influenced cluster composition, though unavoidable for practical reasons. This was mitigated by site selection based on several years of prior malaria data provided by the RCAF. However, past rates cannot be relied on to predict actual transmission, and did not entirely do so here.

## Conclusions

The all-encompassing approach to malaria elimination described here among mobile populations on the Cambodian-Thai border demonstrated that a chemoprophylaxis strategy (MMP) was more effective than a reactive-screen-and-treat approach (FSAT). However, FSAT intervention was improved by the addition of ITUs. The effects of MMP were longer lasting after the intervention was completed. This was true for both *Pf* and *Pv*, though in this area of MDR *Pf*, follow-up surveillance and retreatment were necessary for 30% of volunteers treated initially with DP. A weekly dose of 22.5mg primaquine was safe, well-tolerated, and appeared to potentiate DP prophylactic effects particularly against Pv. Individual contributions of weekly PQ could not be established. FSAT was not effective in reducing *Pv* prevalence. Insecticide-treated clothing appeared to provide additional benefit with the FSAT but not MMP approach. The MMP approach may be more easily scaled to eliminate malaria given reduced need for diagnostic assessments. The study informs a number of outstanding malaria elimination questions, and highlights regional militaries as a potent force for malaria elimination in the GMS.

The study provided valuable insights into the ongoing fight against malaria in Cambodia and underscores the importance of multifaceted approaches tailored to local epidemiology. Cambodia’s National Malaria Program can make significant strides towards malaria elimination in the future by integrating community engagement and empowerment, targeted chemoprophylaxis, vector control monitoring and implementation of ITUs for military populations, and evaluation into a comprehensive, adaptive approach.

## Data Availability

All data produced in the present study are available upon reasonable request to the authors

## Acknowledgements

We are grateful to the study participants and communities who participated in and supported this study. We also thank the Royal Cambodian Armed Forces for their support.

## Disclaimer

The views expressed in this article are those of the authors and do not reflect the official policy of the US Department of the Army, US Department of Defense, the US Government or the Royal Government of Cambodia.

## Author contributions

Study design: DLS, JEM, PS, JTL, SS, WK, PG

Data collection: All

Data analysis: MW, SC, TB, SS, PG, DLS, WK, TK, PG

Data interpretation and study conclusions: TB, MW, SC, SS, PG, JTL, DLS, JEM, MS, PV

Writing manuscript: MW, DLS, SC,

Editing manuscript: DLS, SC, SS, JEM, JTL, MS

All authors read and approved the final manuscript.

## Availability of data and materials

The dataset supporting the conclusions of this article is included within the article and its additional files.

## Declarations

### Ethics approval and consent to participate

This study was approved by the Institutional Review Board at the Walter Reed Army Institute of Research and the Cambodian National Ethics Committee on Human Research. All study participants provided written study consent to participate.

### Consent for publication

All study subjects agreed to publication of the study results in the medical literature without their identity at consent prior to participation.

## Supplemental Material

**Supplemental Table 1.** Withdrawals (W) and losses to follow-up (LTF) over time by treatment group and total volunteers (TOT).

| Time | MMP W | MMP LTF | MMP ToT | MMP % | FSAT W | FSAT LTF | FSAT ToT | FSAT % | TOT W | TOT LTF | TOT | ToT % |
| --- | --- | --- | --- | --- | --- | --- | --- | --- | --- | --- | --- | --- |
| D1-30 | 7 | 1 | 8 | 1.5% | 5 | 1 | 6 | 1.2% | 12 | 2 | 14 | 1.3% |
| D31-60 | 12 | 0 | 12 | 2.2% | 11 | 4 | 15 | 2.9% | 23 | 4 | 27 | 2.6% |
| D61-90 | 4 | 1 | 5 | 0.9% | 9 | 8 | 17 | 3.3% | 13 | 9 | 22 | 2.1% |
| D91-180 | 19 | 6 | 25 | 4.7% | 25 | 12 | 37 | 7.2% | 44 | 18 | 62 | 5.9% |
| Total | 42 | 8 | 50 | 9.3% | 50 | 25 | 75 | 14.5% | 92 | 33 | 125 | 11.9% |

**Supplemental Table 2.**
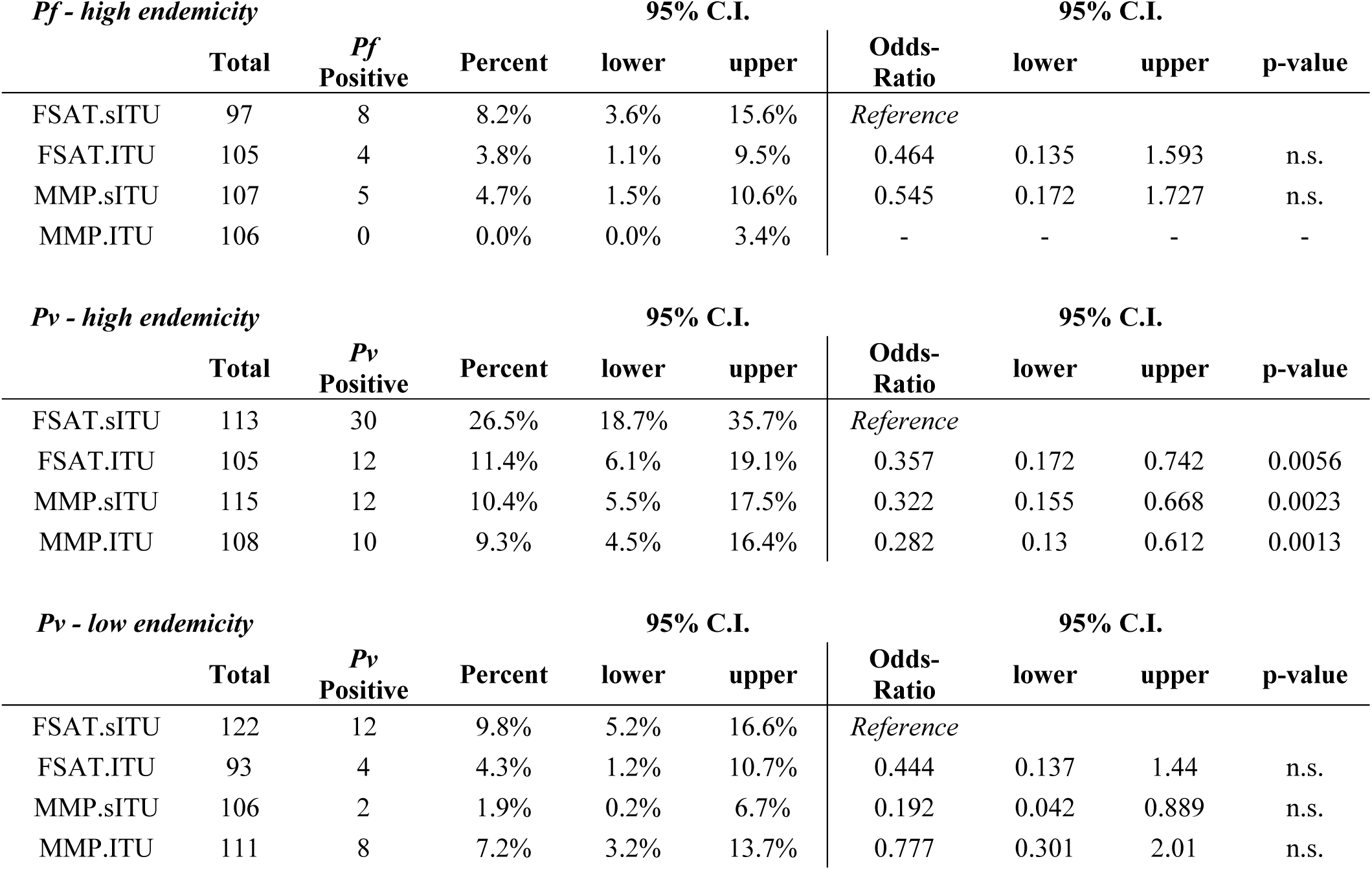
Summary logistic regression analysis results. All comparisons are made with respect to FSAT randomized to sham-treated uniforms (FSAT+sITU). In high Pf-endemicity clusters, both FSAT with treated uniforms and MMP with untreated uniforms had 2-fold less risk for malaria infection (ns). In high Pv endemicity clusters, FSAT with treated uniforms or MMP with or without treated uniforms had a roughly 3-fold significant decrease (95% CI of 1.5-fold to 6.5-fold) in malaria risk (p < 0.01). There was a trend toward decrease in Pv risk in low endemicity clusters associated with treated uniforms or MMP (ns).

### Recurrent *Pv* infection during study follow-up

Nearly all cases of malaria recurrence (26/30, 86%) after the first monthly follow-up were due to *Pv* malaria (**Fig 3C and D**). There were 13 *Pv* recurrences in FSAT treatment group and 13 cases in MMP. These *Pv* recurrences may represent relapses or new infections. Logistic regression revealed that recurrence of *Pv* in MMP was associated with diagnosis of *Pv* on enrollment (p<0.001) and lower weekly PQ dose (<5mg/kg/total dose; p<0.001). Among the volunteers with at least one episode of *Pv* malaria, the cumulative risk of *Pv* recurrence (Kaplan-Meier analysis) was 8% in MMP arm and 17% in FSAT within 150 days of follow up (Supplemental Figure 1A). There were three notable distinctions between *Pv* recurrences in the MMP and FSAT treatment groups. First, in the MMP group, 100% (13/13) of cases of *Pv* recurrence were in individuals that tested positive for *Pv* at enrollment which was significantly higher than the 31% (4/13) of cases in FSAT (p < 0.001). Second, in the FSAT treatment group, subjects wearing sITUs were approximately three times as likely to experience a *Pv* recurrence compared to subjects wearing ITUs (p < 0.01), while in the MMP group there was no significant difference with respect to uniform. Third, the mean interval between the first and second *Pv* infection was significantly longer for MMP, at 166 days (s.d. 23 days) than for FSAT, at 108 days (s.d. 31 days) (p < 10^-4^

**Supplemental Figure 1.**
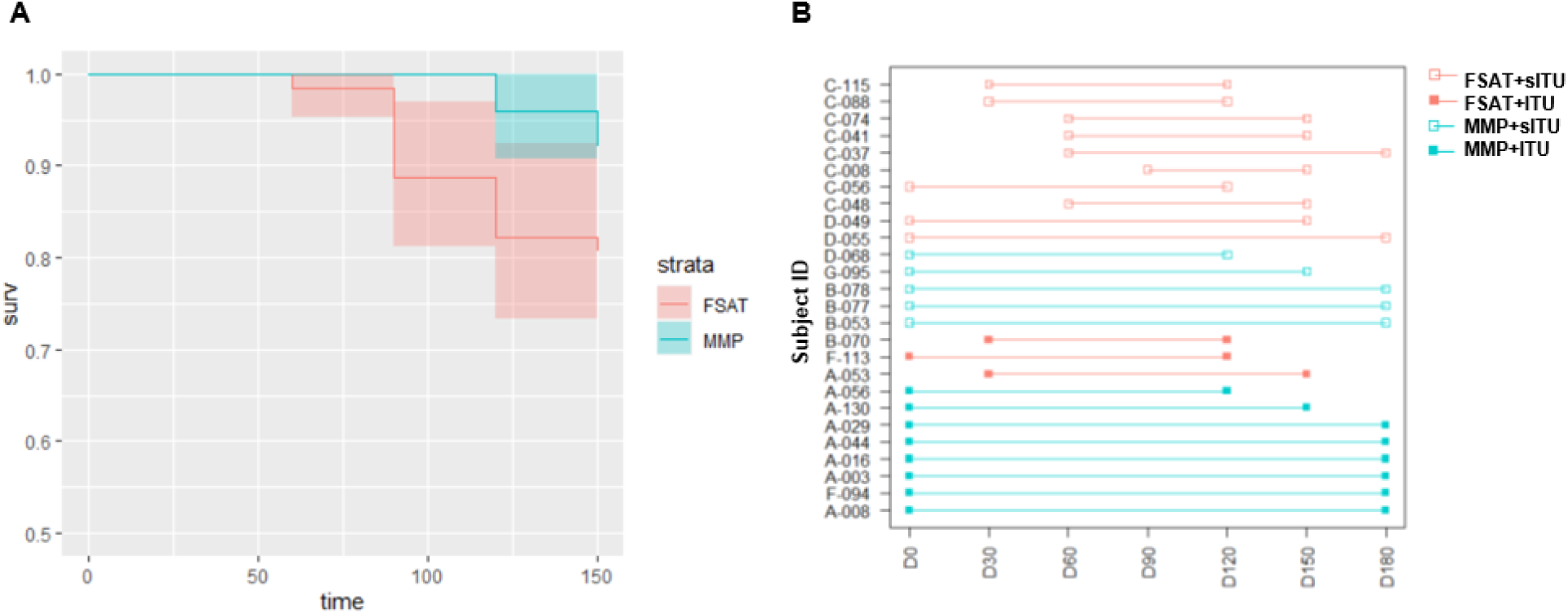
Recurrent *Pv* infections in subjects with at least one *Pv* infection. A) Survival curve to a second *Pv* infection from the first *Pv* infection for all subjects with at least one documented *Pv* infection in the FSAT (red) and MMP (blue) treatment groups. B) Time interval between observing the first and second Pv infection for all subjects that had two documented *Pv* infections in the study. MMP (blue) and FSAT (red) treatment groups are shown as well sITU (open symbol) and ITU (closed symbol).

